# Combined multi-modal assessment of glaucomatous damage with electroretinography and optical coherence tomography/angiography

**DOI:** 10.1101/2020.07.05.20145714

**Authors:** Khaldoon O. Al-Nosairy, Gokulraj Prabhakaran, Konstantinos Pappelis, Hagen Thieme, Michael B. Hoffmann

## Abstract

**Purpose:** To compare the diagnostic performance and to evaluate the interrelationship of electroretinographical and structural and vascular measures in glaucoma.

**Methods:** For 14 eyes of 14 healthy controls and 15 eyes of 12 patients with glaucoma ranging from preperimetric to advanced stages OCT, OCT-A and electrophysiological measures [multifocal photopic negative response ratio (mfPhNR) and steady state pattern electroretinogram (ssPERG)] were applied to assess changes in retinal structure, microvasculature, and function, respectively. The diagnostic performance was assessed via area-under-curve (AUC) measures obtained from ROC analyses. The interrelation of the different measures was assessed with correlation analyses.

**Results:** mfPhNR and ssPERG amplitudes, parafoveal (pfVD) and peripapillary vessel density (pVD), macular ganglion cell inner plexiform layer thickness (mGCIPL) and peripapillary retinal nerve fibre layer thickness (pRNFL) were significantly reduced in glaucoma. The AUC for mfPhNR was highest among diagnostic modalities (AUC: 0.88, 95%-CI: 0.75-1.0, P< 0.001), albeit not statistically different from that for macular (mGCIPL: 0.76, 0.58-0.94, P< 0.05; pfVD: 0.81, .65-.97, P< 0.01) or peripapillary imaging (pRNFL: 0.85, 0.70-1.0, P< 0.01; pVD: 0.82, 0.68-0.97, P < 0.01). Combined functional/vascular measures yielded the highest AUC (mfPhNR-pfVD: 0.94, 0.85-1.0, P<0.001). The functional/structural measure correlation (mfPhNR-mGCIPL correlation coefficient (r_s_): 0.58, P = 0.001; mfPhNR-pRNFL r_s_: 0.66, P < 0.0001) was stronger than the functional-vascular correlation (mfPhNR-pfVD r_s_: 0.29, P = 0.13; mfPhNR-pVD r_s_: 0.54, P = 0.003).

**Conclusions:** The combination of ERG measures and OCT-A improved diagnostic performance in glaucoma. Combing ERG, structural and OCT-A parameters provides an enhanced understanding of the pathophysiology of glaucoma.

## INTRODUCTION

Glaucoma is a progressive optic neuropathy characterized by the loss of retinal ganglion cells (RGCs) and eventually visual field (VF) defects (Kwon et al. 2009). Damage to RGCs is attributed to an increase in intraocular pressure (IOP) (mechanical theory) or primary vascular dysfunction (vascular theory) (Flammer 1994, Flammer et al. 2002, Halpern & Grosskreutz 2002, Mansouri 2016). Elevated IOP is an important risk factor for the development of primary open angle glaucoma (POAG) (Bahrami 2006), the most prevalent glaucoma type (Tham et al. 2014), while vascular dysfunction might be particularly critical for normal tension glaucoma (NTG) (Mansouri 2016). However, vascular changes were also proposed for POAG (Bonomi et al. 2000, Mroczkowska et al. 2013, Salowe et al. 2015). Surrogate measures in clinical practice to estimate glaucomatous damage are macular ganglion cell inner plexiform layer (mGCIPL) and peripapillary retinal nerve fiber layer (pRNFL) thickness measures obtain via optical coherence tomography (OCT) (Kim et al. 2017, Mwanza et al. 2012, Oddone et al. 2016); however, conventional structural OCT assessment does not enable the quantification of vascular changes in glaucoma (Gao et al. 2016). Using the OCT platform for 3-D angiography allows for optical coherence tomography angiography (OCT-A), a recent innovation in imaging technology. In fact, OCT-A has opened the possibility of non-invasive evaluations of retina and optic nerve vasculature in glaucoma (Akil et al. 2018, Jia et al. 2012, Kim et al. 2017, Penteado et al. 2018, Yarmohammadi et al. 2016) to further our understanding of the underlying pathophysiology and to improve glaucoma detection. Vessel density parameters of macular and peripapillary areas measured with OCT-A have a similar diagnostic performance as retinal thickness measures obtained with conventional OCT (reviewed in (Van Melkebeke et al. 2018)). In fact, vessel density measures of OCT-A were strongly correlated with both structural OCT measures and functional indices (standard automated perimetry; SAP) (Van Melkebeke et al. 2018).

Although it is well known that OCT-A correlates with visual field sensitivities (Ghahari et al. 2019, Liu et al. 2015, Yarmohammadi et al. 2016), there is limited information of OCT-A measures correlation with direct measures of retinal ganglion cell function. This gap can be filled by combining OCT-A parameters with electroretinographic (ERG) measures. Two ERG-based approaches provide sensitive information about the pathophysiology of glaucoma damage (Wilsey & Fortune 2016), i.e. pattern ERG (PERG) and the photopic negative responses (PhNR). They are therefore of paramount importance for the objective assessment of retinal function in glaucoma. The PERG is an established method with promising outcomes for glaucoma diagnosis (Bode, Jehle & Bach 2011, Preiser et al. 2013). The PhNR is a more recent development to quantify glaucomatous damage (Kirkiewicz, Lubiński & Penkala 2016, Preiser et al. 2013, Viswanathan et al. 2001), which has been applied in a conventional manner and in combination with the multifocal stimulation technique (Sutter 2001), i.e., mfPhNR (Kaneko et al. 2015, Kato et al. 2015, Van Alstine & Viswanathan 2017).

Taken together, a combined approach of structural, vascular and functional assessment of glaucomatous retinal damage employing OCT, OCT-A and PERG/mfPhNR is of great promise to uncover the interrelationship between the different components of ocular damage in glaucoma and to shed light on the underlying patho-mechanisms. A recent study (Honda et al. 2019) demonstrated that in NTG the PhNR amplitude was correlated with macular vessel density and concluded that vascular changes might precede structural measures in early NTG. We aimed to assess such relationships for POAG. For this purpose, we correlated two types of ERG methods (PERG and mfPhNR) vs structural (OCT based) and vascular (OCT-A based) changes of macular and peripapillary areas. This multimodal approach opens a window to assess how these structural, vascular and functional measures of retinal damage are linked to peripapillary and macular damage sites, and to each other. The aim of the present study was twofold: (i) to compare the diagnostic performance of individual measures and of combined measures of ERG and structural or vascular parameters; (ii) to elucidate the interrelation of ERG measures of retinal ganglion cell function with structural and vascular parameters and their association with macular and peripapillary sites. We found ERG measures (PERG/mfPhNR) to be more strongly correlated with structural (especially peripapillary) measures and less with vascular measures. Combined assessments of multifocal photopic negative responses (mfPhNR) and parafoveal vessel density measures (pfVD) improved the diagnostic power of glaucoma detection.

## METHODS

### Participants

12 glaucoma patients and 14 age-matched healthy controls were included in this observational study after giving written consent to participate in the study. ERG data of the study participants were part of another study (Al-Nosairy, Thieme & Hoffmann 2020). The procedures followed the tenets of the declaration of Helsinki and the protocol was approved of by the ethical committee of the Otto-von-Guericke University of Magdeburg, Germany. The study was performed in the ophthalmological department of the Otto-von-Guericke University Hospital, Magdeburg. All participants underwent best corrected visual acuity testing for far (BCVA) and near, visual field testing, OCT/-A measurements, and an ophthalmic examination.

#### Healthy controls

14 eyes of 14 subjects (mean age ±, standard error of mean (SE): 50.2 years, 3.8) with BCVA ≥ 1 were included in the study.

#### Glaucoma-group

15 eyes of 12 patients (mean age ±, SE: 55.3 years, 3.7; no age difference to control group (P = 0.35; t-Test)), with open angle glaucoma were enrolled in this study. The group comprised 7 preperimetric and 8 perimetric glaucomatous eyes. The 7 preperimetric glaucoma patients with an open anterior chamber, had a glaucomatous optic disc damage defined via a vertical cup-to-disc ratio ≥ 0.7, a retinal fiber layer defect or a local notching of the rim. The 8 perimetric glaucoma eyes had glaucomatous visual field defects manifested as a cluster of 3 or more non-edge points all depressed on the pattern deviation plot < 5% and one of which depressed < 1% or abnormal corrected pattern standard deviation < 5% on Humphrey Swedish interactive threshold algorithm 24-2 (SITA fast) (Anderson & Patella 1999).

Exclusion criteria were any systemic diseases, ocular diseases or surgeries that might affect electrophysiological recordings except cataract surgery and, in the glaucoma group, glaucoma surgery or BCVA < 0.8 (Bach & Mathieu 2004) and refractive error exceeding ±5 D or astigmatism > 2 D. Insufficient quality of OCT images was also an exclusion criteria. An overview of participants’ characteristics is given in Table 1.

**Table 1.**
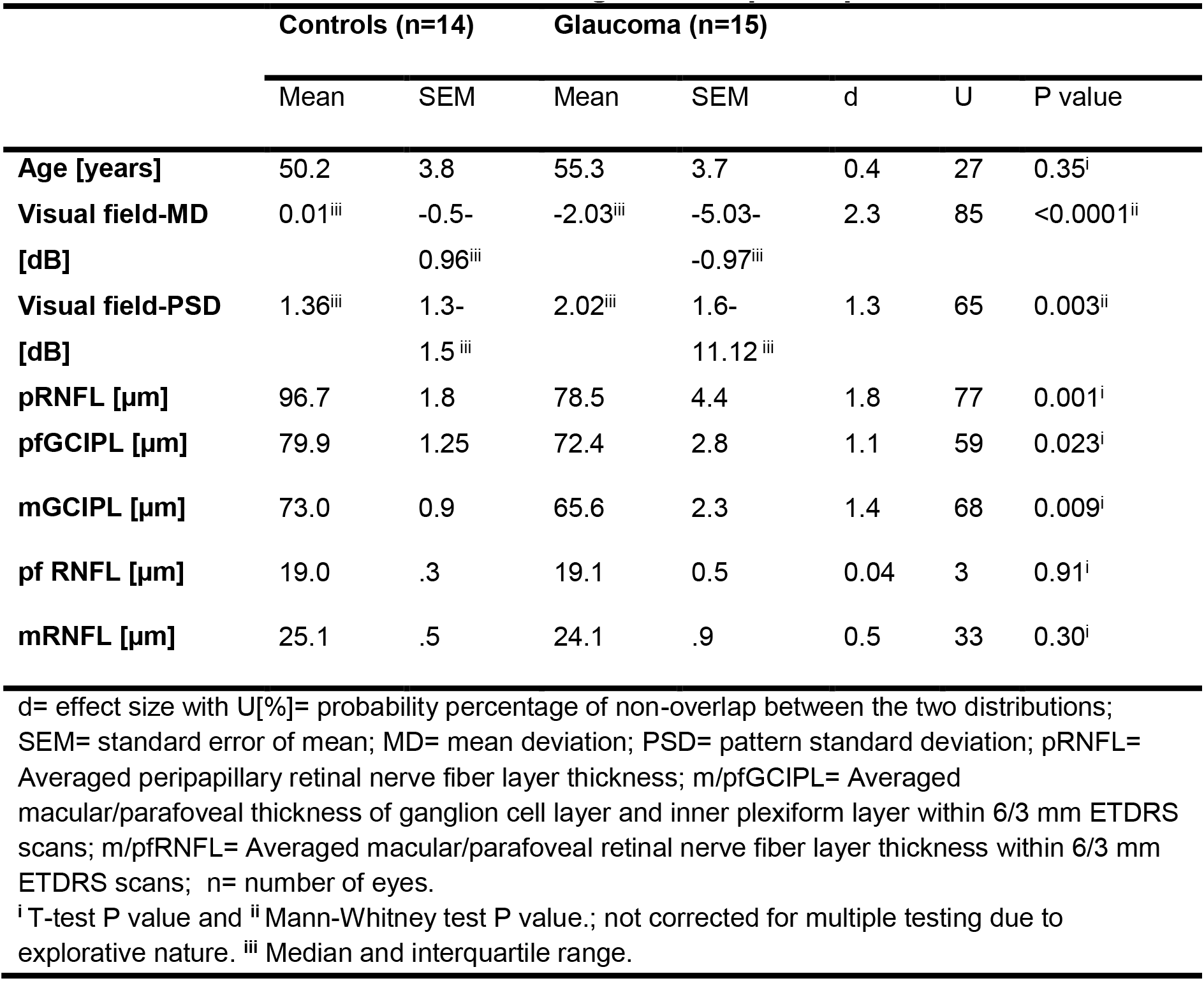
Characteristics of normal and glaucoma participants.

### Visual field testing

Visual field sensitivities were assessed using the Swedish Interactive Threshold Algorithm 24-2 protocol (SITA-Fast) of the Humphrey Field Analyzer 3 (Carl Zeiss Meditec AG, Jena, Germany).

### OCT Angiography acquisition and image analysis

OCT images were acquired using the Spectralis HRA+OCT equipped with the Angiography, the Glaucoma and the Flex Module. Both eyes were scanned for macula and disc scans. OCT angiographical images and structural measures were then exported for further analysis.

#### Structural measures

Averaged macular retinal nerve fiber layer thickness and ganglion cell inner plexiform layer (GCIPL) thickness were assessed inside the 3 mm (pfRNFL and pfGCIPL, respectively) and 6 mm (mRNFL and mGCIPL, respectively) rings of ETDRS scan and exported for further analysis (Figure 1 I & J). The averaged peripapillary retinal nerve fiber layer thickness (pRNFL) was calculated within 3.4 mm circle around the disc. Global indices of macula and peripapillary structure measures, i.e. mGCIPL and pRNFL thickness, respectively, were compared to other parameters.

**Figure 1.**
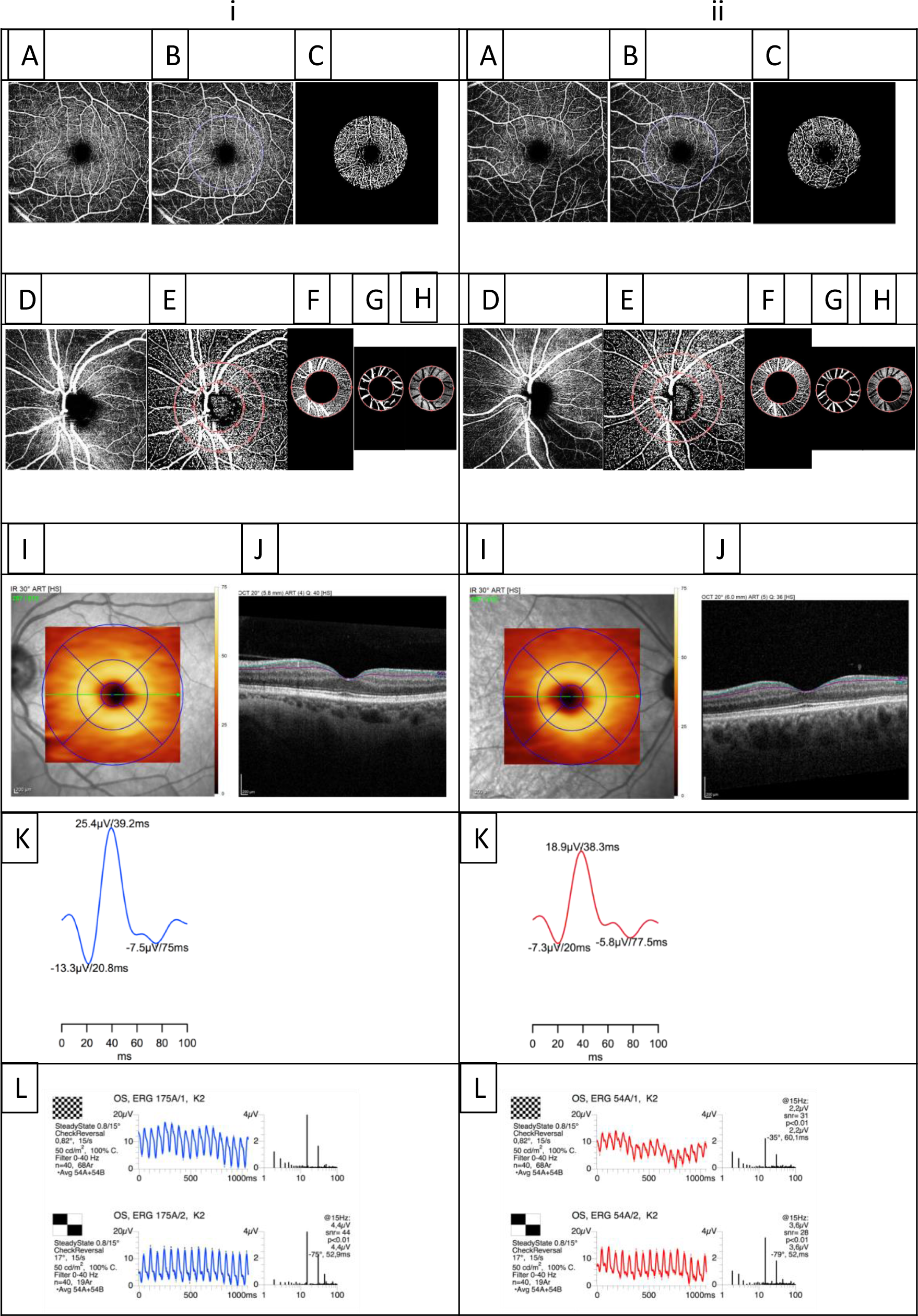
(A) OCT angiography image of the parafovea analyzing superficial vascular plexus (SVP) of (i) a representative control’s and (ii) a glaucoma participant’s left eye. In (B) off-line post-processed images (see text for details) are depicted, where (B) the region of interest (ROI) is delineated and (C) the ROI is used for subsequent analyses. (D) OCT-A of the peripapillary area extracting the superficial vascular complex of (i) a control and (ii) a glaucoma participant. In (E-F) off-line processed images are depicted, where (E) is an image of the disc with delineation of ROI, (F) ROI of disc selected, (G) exclusion of big vessels from the analyzed area (H). In (I) the ETDRS scans of the macula are depicted, with a visualization of the 1, 3, 6 mm circles used for the analyses. In (J) a macular OCT image is shown with the ganglion cell layer embraced between the lines. (K) Averaged mfPhNR trace with the 1^st^ negativity, i.e., a wave, the 1^st^ positivity, i.e., b wave, and the 2^nd^ negativity, i.e. the mfPhNR component. (L) ssPERG to 0.8 checksize (upper panel) and 15° checksize stimuli (lower panel) together with the frequency plot with the dominant response at the stimulation frequency, i.e., 15 Hz, and the corresponding P values of each response.

#### Angiographical measures

Spectralis OCT-A enables distinctive mapping of three vascular layers of the retina, superficial vascular plexus (SVP), intermediate capillary plexus (ICP) and deep capillary plexus (DCP) (Hosari et al. 2020). We focused our analysis on the SVP layer that nourishes macular GCLIPL (mGCIPL) and peripapillary superficial vascular complex (SVC) layer, which includes the peripapillary radial capillaries supplying the pRNFL (Campbell et al. 2017, Iwasaki & Inomata 1986). OCT-A images were exported in the form of transverse analysis from the Heidelberg Engineering interface. High speed scans (20°) were used and 768 x 768 pixel images were utilized. SVP (Figure 1A), ICP and DCP of parafovea were evaluated. Each layer was analyzed separately with a MATLAB-script (Pappelis & Jansonius 2019) described below. Only the peripapillary superficial vascular complex (SVC) of the peripapillary area perfusion was evaluated with the current script (Figure 1D).

With the MATLAB-based script (Pappelis & Jansonius 2019) used for analysis, images were imported and one region of interest (ROI) (see below) was defined after determining the center of macula and disc for macular and disc perfusion quantification by the same investigator, respectively. SVP, ICP and DCP were calculated automatically, once the ROI center was determined manually by the investigator. Binary images of macula and optic disc were generated and each vessel pixel and tissue pixel were represented as white and black, respectively. A local Otsu threshold (Otsu 1979) to binarize an image was applied to flow- and no-flow signals. The ROI of the macula was a circle with 3 mm diameter centered on the macula (Figure 1 C) and the ROI around the optic disc was a ring shaped with inner and outer radii of 1.03 and 1.84 mm (Figure 1 F). The big blood vessels of the optic nerve head (ONH) images were masked out with a Frangi vesselness filter using eignenvectors of the Hessian filter response of image (Frangi et al. 1998) (Figure 1 G & H). To assess the reproducibility of the applied script, repeated analysis of the same OCT-A images were compared between the OCT-A data of the study population. Pairwise comparisons did not identify significant differences between the 2 iterations of image processing (P > 0.05). Intraclass correlations between both data sets of SVP, ICP and DCP showed excellent agreement of all measures (95% CI of ICC: 0.99-1.0, P < 0.001).

The following parameters were evaluated: (1) Fractal dimension (FD). FD is an index of the branching complexity of the capillary network and ranges from 1 to 2, with a higher FD indicating a greater vessel branching pattern. FD was calculated based on a box-counting technique where the image is subdivided into square boxes of equal sizes and the number of boxes covering a vessel segment is counted. This was repeated for different box sizes. The logarithmized box number was plotted vs logarithmized box size, where the FD equals the slope of the regression line (Masters 2004, Reif et al. 2012). (2) Vessel density (VD) [%]. VD is the percentage of the area occupied by capillaries. The peripapillary parameters of FD and VD were denoted as pFD and pVD to differentiate them from parafovea pfFD and pfVD parameters. Only the parafoveal and peripapilary SVP were compared to other functional and structural parameters. In the literature, the most frequently reported measure of microvasculature in glaucoma is VD. Consequently, we focused on VD, also in Discussion, specifically as we obtained similar findings for FD and VD.

### Visual stimuli, procedure and recordings of mfPhNR and ssPERG

#### Multifocal photopic negative response (mfPhNR)

For mfPhNR recording, VERIS Science 6.4.9d13 (EDI: Electro-Diagnostic Imaging, Redwood City, CA, USA) was used for stimulus delivery and electrophysiological recordings. The stimulus comprised 5 visual field locations covering 48° of visual field with central and four quadrants (0-5° and 5°-48°, respectively). A binary m-sequence of 0 (no flash) and 1 states (flash) was used for stimulation with a length of 2^9^-1 steps and 9 frames (frequency of stimulation: 4.2 Hz). Each step lasted 13.3 ms resulting in total recording time of 61 s. Two mfPhNR blocks were recorded and averaged. A monochrome CRT monitor (MDG403, Philips; P45 phosphor) was used for the stimulus presentation at 75 Hz frame rate and the measurements were inspected in real-time on a separate monitor. In accordance with previous studies, mfPhNR were normalized to b-wave amplitude, both measured from the baseline, defined as the initial 10 ms of the epoch. The resulting mfPhNR ratio was compared between groups. We reported only mfPhNR ratio of the summed response of 5 visual field locations (Figure 1 K), as this was previously identified to be the most accurate measure in mfPhNR-based glaucoma diagnostics (Al-Nosairy, Thieme & Hoffmann 2020).

#### Steady state Pattern ERG (ssPERG)

The EP2000 evoked potential system was used for stimulation, recording and analysis of steady-state PERGs (Bach n.d.) following the PERG-standard of the international society for clinical electrophysiology of vision (ISCEV) (Bach et al. 2013). The stimuli were presented at a frame rate of 75 Hz on a monochrome monitor (MDG403, Philips; P45 phosphor) subtending a visual angle of 62°x49°. A 15 Hz checkerboard pattern stimulus with two check sizes (0.8° and 15°) was used for stimulation (figure 1 L). Following established procedures (Bach & Hoffmann 2008), the PERG ratio is calculated as an amplitude ratio of checksizes 0.8 to 15°.

In separate sessions, mfPhNR and ssPERG were recorded binocularly using active DTL (Dawson, trick Litzkow 1979, Thompson, Drasdo, 1987) electrodes (DTL Electrode ERG, Unimed electrode Supplies, Ltd, UK).

The pupils were dilated only for the mfPhNR recordings. Further details on the procedure and recording, analysis of mfPhNR and ssPERG are given in (Al-Nosairy et al. 2020, Al-Nosairy, Thieme & Hoffmann 2020, Preiser et al. 2013). Only the ssPERG 0.8° amplitude (ssPERG amplitude) and averaged mfPhNR/b-wave ratio (mfPhNR ratio) were compared to the acquired vascular and structural parameters.

### Statistics

mfPhNR ratio (mfPhNR) and ssPERG amplitude were calculated using IGOR (IGOR Pro, WaveMetrics, Portland) and exported to SPSS 26 (Statistical Package for the Social Sciences, IBM), and R statistical system (R Core Team (2013) n.d.) for further analysis. The normality of the data was checked by applying the Shapiro-Walk test. T-tests or Mann-Whitney tests were conducted for cross-modal comparisons between groups and effect sizes of these tests were also reported as a d value and U [%] which represented the probability percentage of non-overlap between two distributions (Fritz, Morris & Richler 2012). Correlations between measures were calculated using Spearman’s coefficient (r_s_) and the 95% confidence interval of the coefficient was calculated using a bootstrapping method. The variances explained by the correlations (r_s_^2^) were also calculated and reported. Receiver operating characteristics analyses were conducted using SPSS to calculate area under curve (AUC) to discriminate between controls and glaucoma. Pairwise comparisons of all measures’ AUCs were assessed to check for any significant difference between them (Hanley & McNeil 1983). P values were corrected for multiple testing with adjusted α-levels (Pα) using the Bonferroni-Holm correction (Holm 1979) where applicable. To verify the reproducibility of the applied MATLAB analysis script, intraclass correlation of analyses between two sets of repeated analysis of the same OCT-A images and 95% confidence intervals (95% CI) were calculated based on absolute-agreement and 2-way mixed-effects model (Koo & Li 2016). MATLAB R2019b (MathWorks, Natick, Massachusetts) was used for OCT-A image processing.

## RESULTS

### Functional and structural parameters vs electrophysiological and vascular measures

#### Electrophysiology

The electrophysiological measures of retinal ganglion cell function showed differential responses between the groups. The mfPhNR ratio was significantly different in glaucoma and the difference between the groups represented 75% of the non-overlapping distribution (d = 1.7, P_≤0.025_ = 0.0002). Similarly, the difference between healthy and glaucoma ssPERG amplitudes was statistically significant (d = 1.1, P_≤0.05_=0.006), for effect sizes see Figure 2 A and B.

**Figure 2.**
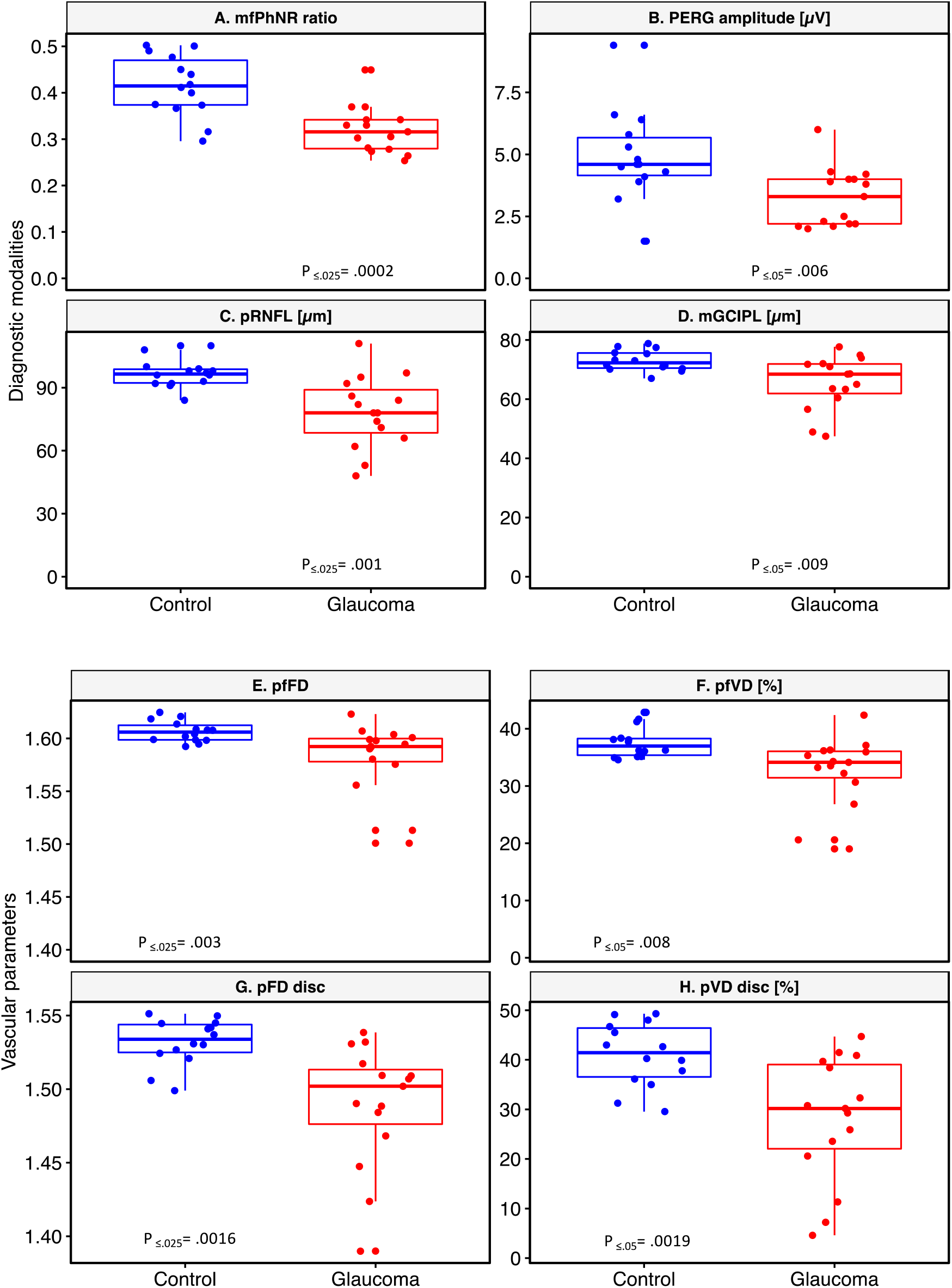
Cross modal comparison of diagnostic performance. (A) Multifocal photopic negative response ratio (mfPhNR), (B) pattern electroretinogram (PERG) amplitude for 0.8° checksize, (C) peripapillary retinal nerve fiber layer thickness in micrometer, and (D) macular ganglion cell layer and inner plexiform layer thickness (mGCIPL). Vascular metrics of (E) parafoveal fractal dimension (pfFD) and (F) parafoveal vessel density (pfVD). Vascular metrics of (G) peripapillary fractal dimension (pFD) and (H) peripapillary vessel density (pVD). Independent t-tests were conducted except for parafoveal FD where Mann-Whitney test was performed (alpha-thresholds corrected for multiple comparisons are shown as subscripts). Panel title specifies the y-axis for each plot.

#### Perimetry

On average, functional measures of glaucoma in terms of VF-MD [dB] and pattern standard deviation (PSD) [dB] were statistically different between the study groups (d = 2.3, P_≤0.025_ < 0.0001 and d = 1.3, P_≤0.05_ = 0.003, respectively; see Table 1).

#### OCT

pRNFL were significantly lower in glaucoma patients with a substantial effect size of 1.8 (P_≤0.025_ = 0.001) (Figure 2 C-D). The mRNFL was not statistically different between the groups (P > 0.05). In contrast, pfGCIPL and mGCIPL thickness were significantly lower in glaucoma (d = 1.1, P_≤0.05_ = 0.023 and d = 1.4, P_≤0.025_ = 0.009, respectively; see Table 1).

#### OCT-angiography

In terms of vascular estimates for the parafoveal ROI, we were particularly interested in the inner retinal layer supplied by SVP (for effect sizes see Figure 3 E-H). Parafoveal FD (pfFD) (d = 1.3, P_≤0.025_ = 0.003) and parafoveal VD (pfVD) (d = 1.1, P_≤0.05_ = 0.008) were significantly reduced in glaucoma. For the peripapillary ROI perfused by SVC, pFD showed a significant decrease (d = 1.7, P_≤0.025_ = 0.0016) as well as pVD (d = 1.5, P_≤0.05_ = 0.0019) in glaucoma patients. It is notable that both ICP and DCP showed significant pfFD and pfVD reductions in glaucoma (P < 0.01) compared to controls (Table 3).

**Figure 3.**
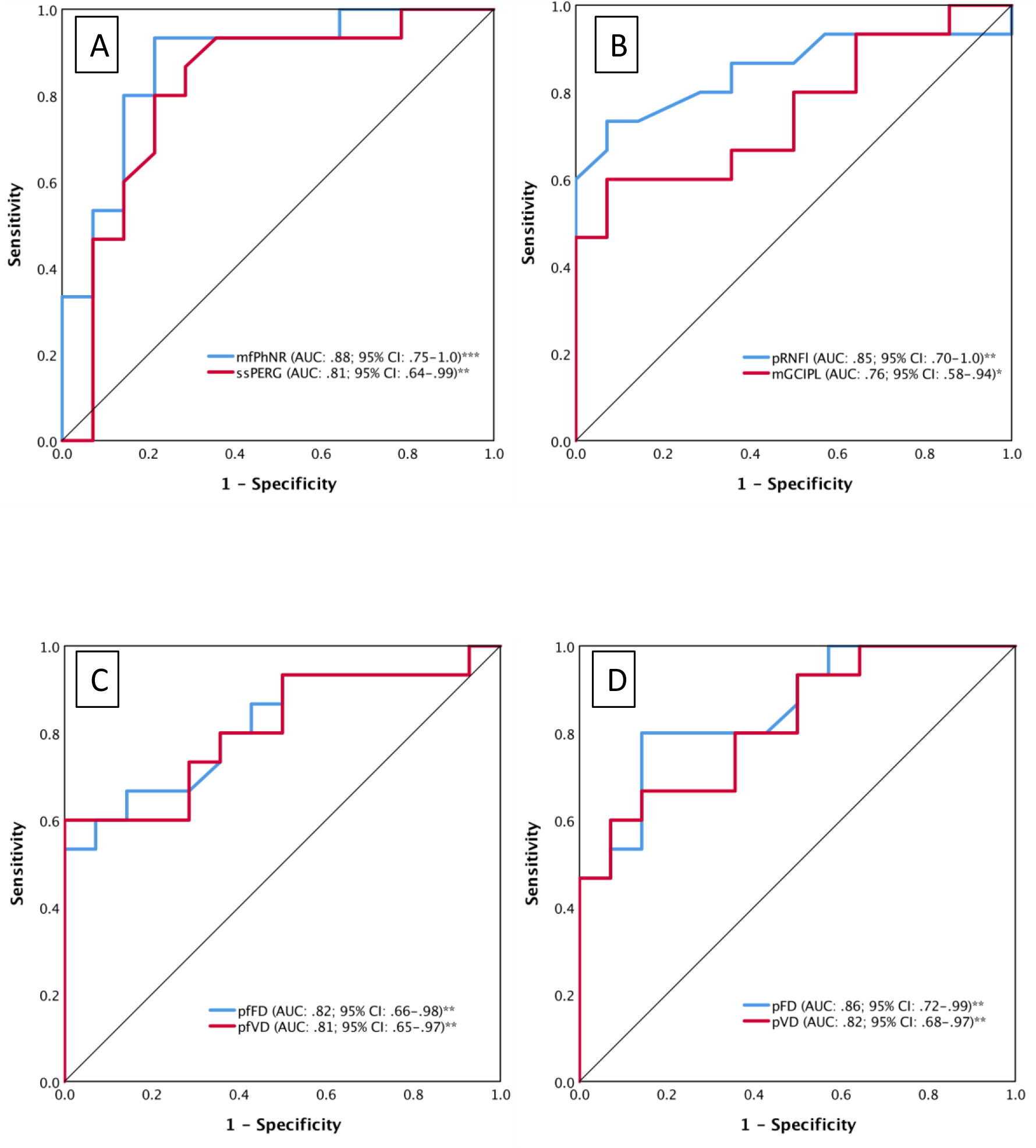
Area under curve (AUC) of receiver operating characteristics (ROC) and AUC 95% confidence intervals (CI). (A) Electrophysiological parameters, multifocal photopic negative response ratio (mfPhNR) and pattern electroretinogram 0.8° amplitude (ssPERG). (B) Structural measures of peripapillary retinal nerve fiber layer thickness (pRNFL) and macular ganglion cell layer and inner plexiform layer thickness (mGCIPL). (C, D) vascular metrics of parafovea, which are (C) parafovea fractal density (pfFD) and vessel density (pfVD) vs peripapillary vascular metrics and (D) peripapillary fractal density (pFD) and peripapillary vessel density (pVD). P values Significance levels are indicated where * indicates P < 0.05, ** indicates P < 0.01 and *** indicates P < 0.001 where the null hypothesis is that true area= 0.5.

**Table 2.**
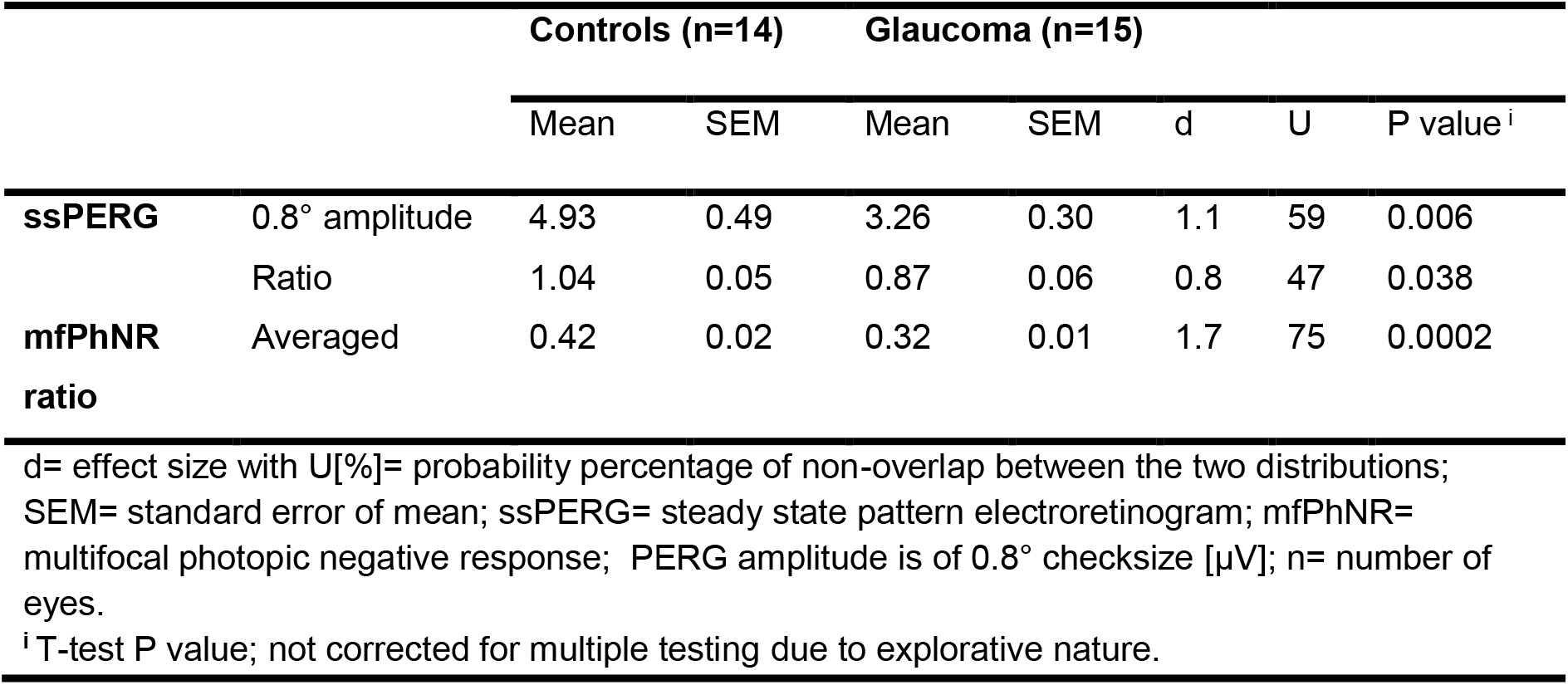
Overview of electrophysiological parameters.

**Table 3.** Overview of vascular parameters.

|  |  |  | Controls (n=14) |  | Glaucoma (n=15) |  | d | U | P value |
| --- | --- | --- | --- | --- | --- | --- | --- | --- | --- |
|  |  |  | Mean | SEM | Mean | SEM |  |  |  |
| **Parafovea** | SVP | pfFD | 1.60<sup>iii</sup> | 1.60-1.61<sup>iii</sup> | 1.59<sup>iii</sup> | 1.58-1.60<sup>iii</sup> | 1.3 | 65 | 0.003<sup>ii</sup> |
|  |  | pfVD | 37.60 | 0.72 | 32.52 | 1.59 | 1.1 | 59 | 0.008<sup>i</sup> |
|  | ICP | pfFD | 1.58<sup>iii</sup> | 1.58-1.59<sup>iii</sup> | 1.57<sup>iii</sup> | 1.56-1.57<sup>iii</sup> | 1.4 | 68 | 0.001<sup>ii</sup> |
|  |  | pfVD | 29.48 | 0.51 | 26.50 | 0.89 | 1.1 | 59 | 0.009<sup>i</sup> |
|  | DCP | pfFD | 1.59 | 0.003 | 1.57 | 0.005 | 1.2 | 62 | 0.0034<sup>i</sup> |
|  |  | pfVD | 31.14 | 0.68 | 27.63 | 0.83 | 1.2 | 62 | 0.0033<sup>i</sup> |
| **Peripapillary disc area** | SVC | pFD | 1.53 | 0.004 | 1.49 | 0.01 | 1.7 | 75 | 0.0016<sup>i</sup> |
|  |  | pVD | 41.01 | 1.73 | 28.06 | 3.27 | 1.5 | 71 | 0.0019<sup>i</sup> |

d= effect size with U[%]= probability percentage of non-overlap between the two distributions;
SEM= standard error of mean; SVP= superficial vascular plexus; ICP= intermediate capillary plexus; DCP= deep capillary plexus; SVC= superficial vascular complex; pf/pFD=
parafoveal/peripapillary fractal dimension; pf/pVD= parafoveal/peripapillary vessel density.
<sup>i</sup> T-test P value and <sup>ii</sup> Mann-Whitney test P value; not corrected for multiple testing. <sup>iii</sup> Median and interquartile range.

### Discriminatory performance of ERG, structural parameters and vascular parameters

In terms of the discriminatory performance between controls and glaucoma, we applied ROC-analyses to compare ERG measures of RGC-function (mfPhNR ratio, ssPERG amplitude), established structural (i.e., mGCLIPL thickness, pRNFL thickness) vascular measures of parafoveal and optic nerve (pfFD and pfVD as well as pFD and pVD). With respect to the ERG measures of RGC-function, there was a, non-significant trend for higher AUC (AUC, 95% CI, P value) for the mfPhNR ratio (0.88, 0.75-1.0, P_≤0.025_ < 0.001) than for the ssPERG amplitude (0.81, 0.64-0.99, P_≤0.05_ = 0.004). Therefore, our further analyses were focused on mfPhNR ratio. With respect to the structural assessment, there was a non-significant trend for higher AUC for pRNFL (0.85, 0.70-1.0, P_≤0.025_ = 0.001) than for mGCIPL (0.76, 0.58-0.94, P_≤0.05_ = 0.018). AUCs for vascular parameters were calculated for pfFD (0.82, 0.66-0.98, P_≤0.025_ = 0.0037) and for pfVD (0.81,0.65-0.97, P_≤0.05_ = 0.005) compared to pFD (0.86, 0.72-0.99, P_<0.025_ = 0.001) and pVD (0.82, 0.68-0.97, P_≤0.05_ = 0.003; see Figure 3). Finally, by conducting pairwise comparisons of ERG measures of RGC-function, structural and vascular AUCs, we found no significant differences (P > 0.05) between these measures indicating a similar and complementary performance in terms of differentiating glaucoma from controls. By testing the combined approach to identify the highest discriminatory performance, mfPhNR-pfVD and mfPhNR-pVD had the highest AUC for the differentiation between glaucoma and controls (AUC: 0.94, 0.91, respectively; P < 0.001).

### Association between ERG, structural parameters and vascular parameters

To elucidate associations between functional and other metrics, we investigated the correlation between vascular estimates of inner layers macula and peripapillary zones vs other structural and ERG measures of RGC-function. Both pfFD and pfVD were strongly correlated with pf/mGCLIPL thickness (P ≤ 0.001; Table 4). Similarly vascular estimates of peripapillary perfusion showed a strong significant association with pRNFL thickness (P ≤ 0.0001). Our ERG measure of RGC-function, the mfPhNR ratio, was strongly correlated with all structural macula and peripapillary disc parameters as well as visual field-MD (P ≤ p.001). ssPERG amplitude was also significantly correlated to pRNFL, mGCIPL and VF-MD (P = 0.003, 0.027 and 0.003, respectively), but not to pfGCIPL (P = 0.09). Out of the vascular measures, the mfPhNR ratio as well as ssPERG amplitude were significantly correlated only with pFD and pVD (P < 0.01; see table 4 and figure 4).

**Table 4.** Correlations between electrophysiological, structural and vascular parameters.

| | | $r_s$, P value, [95% confidence interval of $r_s$]<sup>c</sup> | | | | | | | | |
| --- | --- | --- | --- | --- | --- | --- | --- | --- | --- | --- |
|  |  | Functional |  |  | Structural |  |  | Vascular |  |  |
|  |  | mfPhNR | PERG | VF-MD | pRNFL | pfGCIPL | mGCIPL | pfFD | pfVD | pFD |
| Functional | PERG | 0.60<br>0.001<br>[0.34-0.78] | 1 |  |  |  |  |  |  |  |
|  | VF-MD | 0.76<br><0.0001<br>[0.53-0.89] | 0.53<br>0.003<br>[0.19-0.80] | 1 |  |  |  |  |  |  |
| Structural | pRNFL | 0.66<br>0.0001<br>[0.26-0.90] | 0.53<br>0.003<br>[0.20-0.78] | 0.62<br>0.0003<br>[0.27-0.87] | 1 |  |  |  |  |  |
|  | pfGCIPL | 0.58<br>0.001<br>[0.29-0.77] | 0.32<br>0.09<br>[-.05-0.66] | 0.48<br>0.008<br>[0.07-0.79] | 0.56<br>0.002<br>[0.23-0.78] | 1 |  |  |  |  |
|  | mGCIPL | 0.58<br>0.001<br>[0.28-0.79] | 0.41<br>0.027<br>[0.06-0.70] | 0.51<br>0.005<br>[0.13-0.81] | 0.73<br><0.0001<br>[0.49-0.86] | 0.93<br>0.0001<br>[0.84-0.97] | 1 |  |  |  |
| Vascular | pfFD | .34<br>.07<br>[.001-.62] | 0.24<br>0.20<br>[-.17-0.58] | 0.52<br>0.0004<br>[0.17-0.76] | 0.30<br>0.12<br>[-.11-0.64] | 0.57<br>0.001<br>[0.27-0.76] | 0.57<br>0.001<br>[0.25-0.78] | 1 |  |  |
|  | pfVD | 0.29<br>0.13<br>[-.05-0.58] | 0.21<br>0.27<br>[-.18-0.55] | 0.52<br>0.004<br>[0.20-0.74] | 0.29<br>0.12<br>[-.12-0.63] | 0.56<br>0.001<br>[0.25-0.76] | 0.57<br>0.001<br>[0.25-0.80] | 0.99<br>< 0.0001<br>[0.98-1.0] | 1 |  |
|  | pFD | 0.56<br>0.002<br>[.19-.82] | 0.52<br>0.004<br>[0.19-0.75] | 0.54<br>0.002<br>[0.18-0.79] | 0.80<br>0.0001<br>[0.60-0.92] | 0.49<br>0.007<br>[0.12-0.76] | 0.65<br>0.0001<br>[0.33-0.86] | 0.39<br>0.037<br>[0.04-.067] | 0.37<br>0.046<br>[0.02-0.65] | 1 |
|  | pVD | 0.54<br>0.003<br>[0.14-.082] | 0.49<br>0.007<br>[0.19-0.74] | 0.48<br>0.008<br>[0.08-0.78] | 0.79<br>0.0001<br>[0.57-0.93] | 0.45<br>0.014<br>[0.06-0.73] | 0.64<br>0.0001<br>[0.34-0.84] | 0.35<br>0.07<br>[-.07-0.66] | 0.34<br>0.07<br>[-.07-0.65] | 0.99<br><0.0001<br>[0.98-1.0] |

peripapillary measures= Blue font; macular/parafoveal measures= green fonts.
mfPhNR= multifocal photopic negative response ratio; PERG= steady state pattern electroretinogram of 0.8° check size amplitude, pRNFL= peripapillary retinal nerve fiber layer thickness; pfGCIPL= parafoveal ganglion cell layer and inner plexiform layer thickness within 3 mm ETDRS macular scans; mGCIPL= macular ganglion cell layer and inner plexiform layer thickness within 6 mm ETDRS macular scans; VF-MD= visual field defect mean deviation in decibels [dB]; pfSVP= parafoveal superficial vascular plexus; pSVC= peripapillary superficial vascular complex; pf/pFD= parafoveal/peripapillary fractal dimension; pf/pVD= parafoveal/peripapillary vessel density.
Significance levels of correlation (uncorrected, 2-tailed)
P < 0.01 level (blue), P < 0.05 level (light blue), P > 0.05 level (White)
c Bootstrap results are based on 1000 bootstrap samples.
$r_s$ Spearman's correlation coefficient.

**Figure 4.**
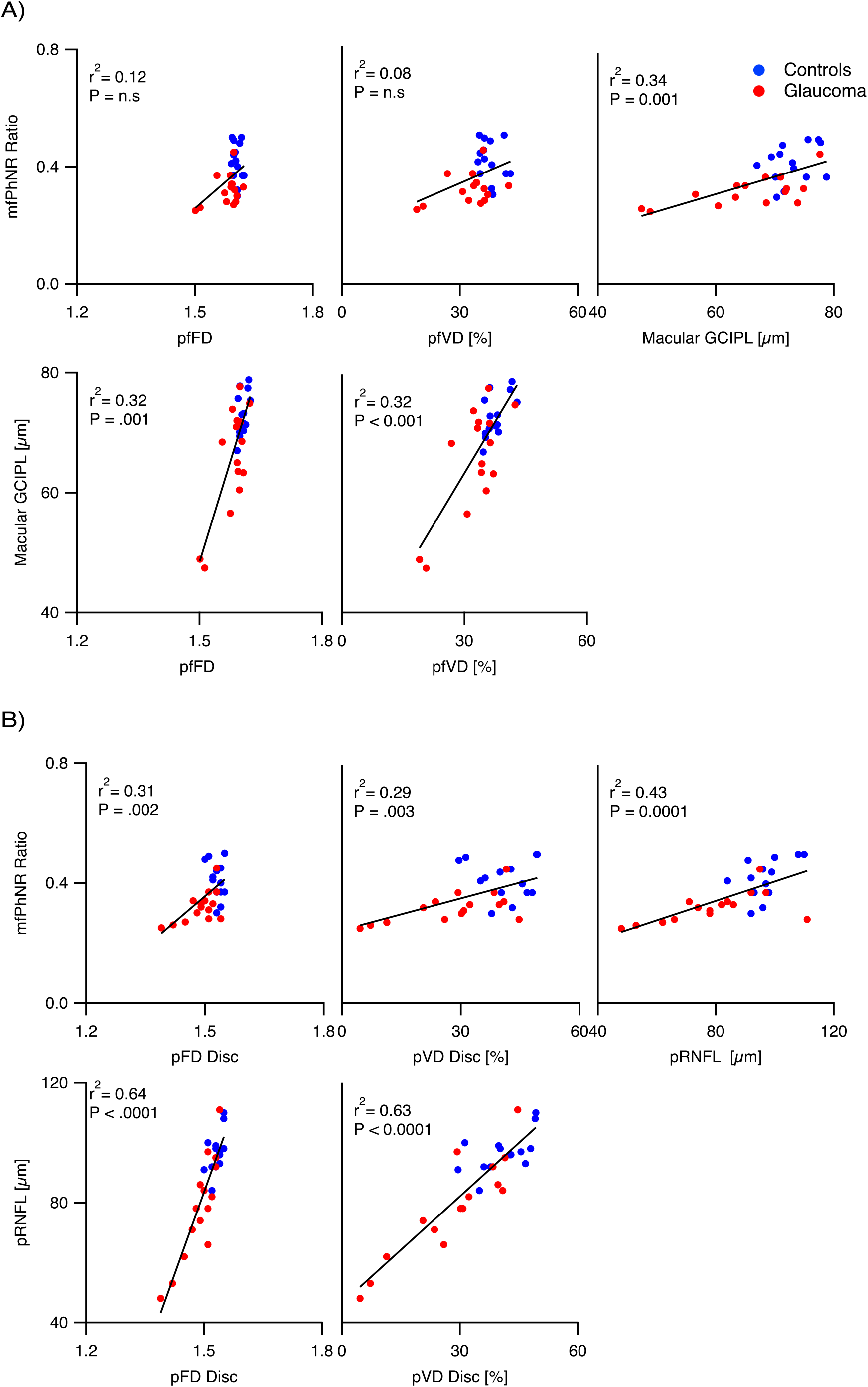
(A) Correlation plots of multifocal photopic negative response ratio (mfPhNR) (upper panel) vs parafoveal fractal density (pfFD) and vessel density (pfVD) and macular ganglion cell layer and inner plexiform layer thickness (mGCIPL) and of mGCIPL (bottom) vs pfFD and pfVD measures. (B) Correlation plots of mfPhNR (top) vs peripapillary perfusion metrics and peripapillary retinal nerve fiber layer thickness (pRNFL) and of pRNFL (bottom) vs peripapillary fractal density (pFD) and vessel density (pVD). r_s_^2^ = coefficient of determination. n.s= non-significant association.

To further elucidate glaucomatous damage mechanisms, we investigated the association between ERG-based functional indices with anatomical indices at damage sites. ERG-based functional measures at the peripapillary site (i.e. mfPhNR-pRNFL r_s_: 0.66, P = 0.0001 and mfPhNR-pVD r_s_: 0.54, P = 0.003) exceeded those at the macular site (i.e. mfPhNR-mGCIPL r_s_: 0.58, P = 0.001 and mfPhNR-pfVD r_s_: 0.29, P = 0.13).

## DISCUSSION

Applying a set of complementary retinal imaging modalities we demonstrated a significant effect of glaucoma on vascular (OCT-A; parafoveal vessel density “pfVD” and fractal dimension “pfFD” and peripapillary pVD and pFD), electrophysiological (mfPhNR and ssPERG amplitude) and structural measures (OCT; mGCIPL/pRNFL). These measures had equivalently high discriminatory performance, which further improved for the combination of the methods. The ERG measures of retinal ganglion cell function were more strongly associated with structural than with vascular measures.

Our findings of significant changes in the ocular microvasculature (VD) in glaucoma support previous studies, that demonstrated glaucomatous changes in the VD of the macular/parafoveal superficial layers (Akil et al. 2018, Chung et al. 2017, Penteado et al. 2018, Rao et al. 2016, Yarmohammadi et al. 2018) and the peripapillary area (Chung et al. 2017, Liu et al. 2015, Rao et al. 2016, Scripsema et al. 2016, Yarmohammadi et al. 2018). Further, they are in agreement with investigations that demonstrated glaucoma associated changes in mfPhNR and PERG (Al-Nosairy, Thieme & Hoffmann 2020, Bode, Jehle & Bach 2011, Preiser et al. 2013) and mGCIPL and pRNFL (Kim et al. 2017, Mwanza et al. 2012, Oddone et al. 2016). We considerably extended these studies by demonstrating an association between ERG-based functional and anatomical indices as well as an enhanced diagnostic efficacy of combined ERG-based functional indices with vascular indices.

### Cross-modal comparison of glaucoma detection

In order to assess the benefit of any of the applied modalities for glaucoma detection, we conducted ROC analyses and compared their outcome measures, i.e. AUC. The only previous cross-modal study addressing this for early glaucoma detection (Kurysheva et al. 2018), demonstrated ssPERG to have a higher performance (AUC = 0.92) than whole image VD in macula and disc (AUC= 0.80 and 0.74, respectively) and ganglion cell complex thickness (AUC= 0.74). In the current study, the highest discrimination performance was observed for the mfPhNR (not tested in (Kurysheva et al. 2018); AUC= 0.88), albeit not being significantly different from other measures’ AUCs. Subsequently, we investigated the effect of combining ERG measures of RGC-function with structural or vascular measures. In fact, the combination of mfPhNR with pfVD and pVD AUCs yielded the highest AUC (0.94 and 0.91, respectively; P < 0.001), indicating an improvement of diagnostic performance. In addition to its relevance for glaucoma diagnosis, the improved performance for the combined assessment with these two modalities might also suggest that the ERG measures of RGC-function and OCT-A measures reflect distinctive glaucomatous damage mechanisms within the retina. It should be noted, however, that, as an alternative, the enhancement might also be due to decreasing the effect of noise by pooling data from different modalities.

### Association of ERG, structural, and vascular measures in glaucoma

Given the relation of vascular changes with glaucoma, it is currently still unresolved, whether these are secondary or primary events associated with RGCs damage (Mansouri 2016). Previous OCT-A studies are inconclusive as they found structural changes either to precede (Akagi et al. 2016, Kim et al. 2017, Lee et al. 2017) or succeed (Honda et al. 2019, Shoji et al. 2017) vascular changes in glaucoma. We investigated the interrelation of these measures with the sensitive measures of RGC-function, mfPhNR and ssPERG amplitude (Al-Nosairy, Thieme & Hoffmann 2020, Bode, Jehle & Bach 2011, Preiser et al. 2013), in order to elucidate glaucomatous damage mechanisms. For this purpose, we compared the association of retinal ganglion cell dysfunction with specific changes (i) in fundus anatomy, i.e., microvasculature (OCT-A) and retinal structure (OCT), and (ii) at damage sites, i.e. macular and peripapillary sites: (i) Fundus anatomy. We reported a stronger correlation of RGC-function (mfPhNR/ssPERG) with retinal structure (r_s_ ≤ 0.66) than with the microvasculature (r_s_ ≤ 0.56). In contrast, for NTG the reverse pattern was recently reported (Honda et al. 2019), i.e. a stronger association of PhNR with measures of macular/parafoveal microvasculature (r ≤ 0.42). Taken together, these findings support the current view that NTG is more strongly associated with vascular damage mechanisms than POAG. (ii) Damage sites. The measures of RGC-function were more strongly associated with peripapillary than with macular structural and vascular measures (mfPhNR with pRNFL and pVD r_s_: 0.66 and 0.54, respectively; mfPhNR with mGCIPL and pfVD r_s_: 0.58 and 0.29, respectively). This suggests that damage mechanisms exert their action preferentially at the peripapillary zone. It must be noted, however, that in the present study glaucomatous damage ranged from preperimetric to advanced glaucoma, such that e.g. early stage changes of the macula (Hood et al. 2013, Kim et al. 2017, Kim, Jeoung & Park 2017) might not have been relevant.

In conclusion, combining ERG and OCT-A measures may improve the assessment and eventually the management of glaucoma. Follow-up studies comparing the effects of glaucoma on retinal electrophysiology, microvasculature, and structure with larger sample sizes and employing longitudinal designs are of promise to further explore the pathophysiology of glaucoma.

## Data Availability

Data available upon reasonable request.

## Acknowledgements

This work was supported by European Union’s Horizon 2020 research and innovation programme under the Marie Sklodowska-Curie grant agreement (No. 675033) and by funding of the German research foundation (DFG; HO2002/20-1) to MBH.

